# Investigating the causal network of dementia by employing a causal discovery approach combined with natural language processing models

**DOI:** 10.1101/2024.11.05.24316664

**Authors:** Xinzhu Yu, Artitaya Lophatananon, Vivien Holmes, Kenneth R. Muir, Hui Guo

## Abstract

**INTRODUCTION:** Comprehensively studying modifiable risk factors altogether to explore how they contribute to dementia mechanism is imperative for effective interventions.

**METHODS:** This study utilized natural language processing (NLP) models to pre-select candidate risk factors of dementia from 5,505 variables in the UK Biobank. We then took a holistic machine learning approach, fast causal inference in combination with mixed graphical models, to explore the complex causal mechanisms underlying dementia from 120 imputed variables.

**RESULTS:** The identified causal network highlighted eight risk factors which may directly or indirectly contribute to dementia. In particular, mental disorders due to brain damage and dysfunction and to physical disease were identified as direct causes as well as mediators on the causal pathways to dementia. Evidence for a direct causal impact of phenotypic age on dementia was less pronounced.

**DISCUSSION:** Our study offered valuable insights into the mechanisms of dementia. Beyond direct connections to nerve or brain disorders, the potential direct link with biological age highlights its possible value in dementia management. Moreover, the use of NLP models for variable pre-selection introduced an innovative application to medical research. Our study added weight to the accruing evidence that machine learning has a promising future for exploring complex disease mechanisms from high-dimensional data.

## 1 Background

Dementia is an increasing global health threat, currently affecting over 55 million people worldwide [1]. The number is expected to triple by 2025, posing significant challenges to healthcare [1]. Identification of modifiable causal risk factors that could be the basis for the prevention of dementia is of increasing scientific interest.

Age is the known strongest risk factor for dementia [2], which is however, non-modifiable. The concept of biological age has been introduced as a biomarker designed to capture the aging processes at the biological level more accurately than chronological age [3]. Phenotypic age, calculated from 10 biomarkers, is not only associated with chronological age but also closely linked to various bodily functions and disease risks [4], [5]. These biological age measurements have demonstrated their potential to effectively predict both overall mortality and specific health outcomes like dementia, surpassing the predictive power of chronological age [3].

The Lancet Commission has reported 14 modifiable risk factors for dementia [6]. In a comprehensive meta-analysis of over 800 reviews, Anstey et al. identified 39 dementia-associated risk factors, spanning lifestyle factors such as physical activity and diet, as well as medical conditions like stroke and renal disease [7]. However, the underlying causal mechanisms linking these risk factors to dementia remain unclear. To inform effective disease intervention, it is essential to systematically and objectively examine the potential pathways and interactions through which these factors may contribute to dementia.

In the era of big data [8], causal discovery from high-dimensional observational data can be particularly challenging. It involves identifying causal connections among numerous variables while accounting for many potential confounders. Advanced machine learning techniques have been developed for such purpose, such as Double Machine Learning [9] and frameworks based on Structural Causal Models [10]. However, these approaches typically assume all confounders are observed—a strong assumption that cannot be verified using only observed data. Fast Causal Inference (FCI) [11] is a machine learning causal discovery approach designed to overcome the limitations of assuming that all confounders are observed. FCI operates by conducting a series of conditional independence tests. By identifying patterns of dependency and independence, FCI infers potential causal structures that account for hidden confounders, resulting in a Partial Ancestral Graph (PAG) that represents plausible causal connections consistent with the data [12]. These methods are emerging as powerful tools for efficiently identifying complex data structures, offering new evidence and insights into biological processes and disease etiology [13].

Here, we exploited graphical and machine learning approaches to systematically examine how modifiable risk factors contribute to pathways to dementia collectively and map them out into a network. Additionally, two pre-trained information retrieval (IR) models [14] [15] were employed to select dementia-related factors from a large pool of variables in the UK Biobank, with the aim to evaluate whether natural language processing (NLP) models have the potential to help variable selection from extensive databases.

## 2 Methods

The workflow of the study is represented in Figure 1.

**Figure 1.**
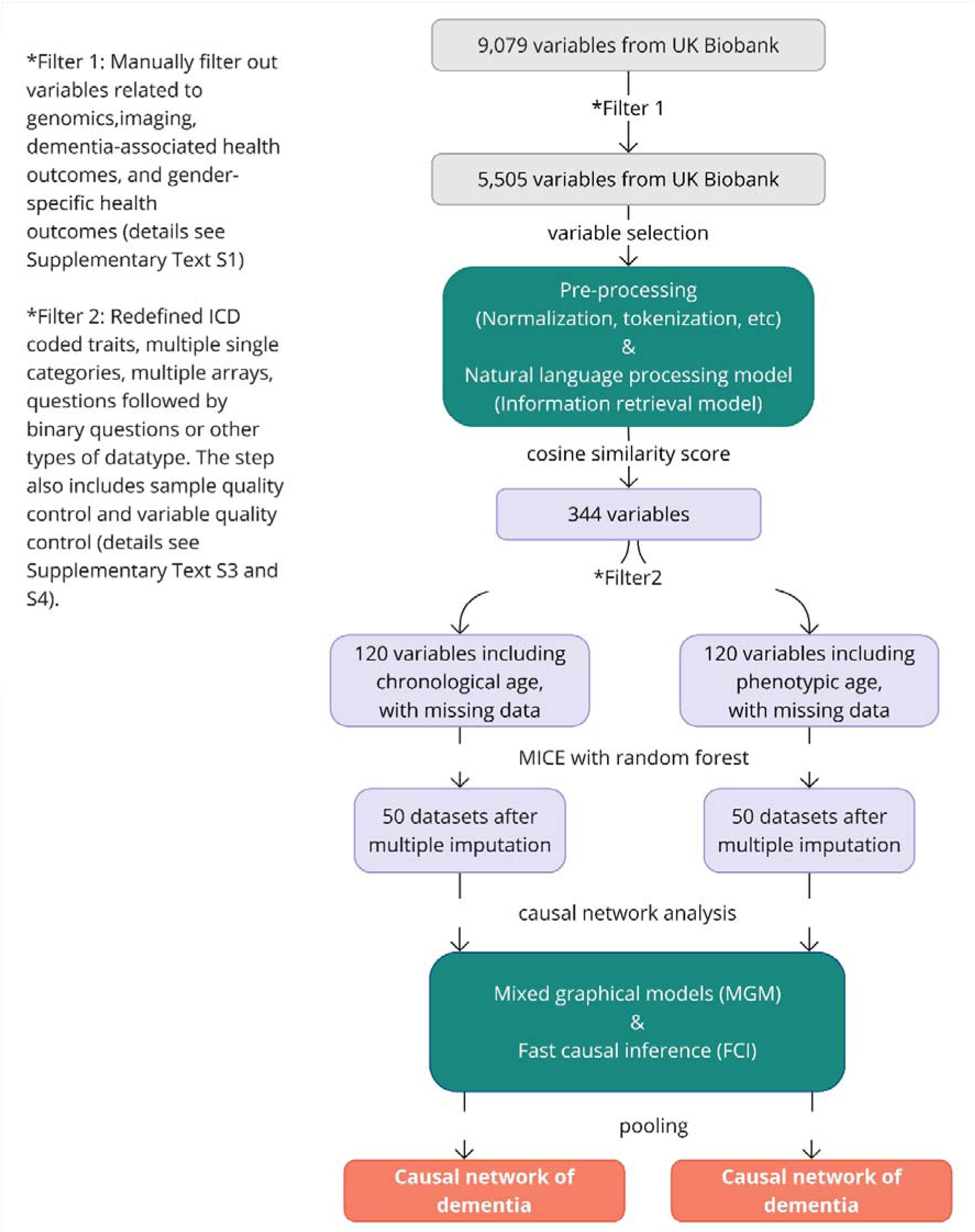
Workflow of the study. This workflow diagram illustrates the pre-processing and analysis steps of the study. Green box 1 details the variable selection step using natural language processing models. The variables initially filtered from the UK Biobank data dictionary are further selected using natural language processing models based on cosine similarity scores. Green Box 2 outlines the causal network analysis steps - Mixed Graphical Models and Fast Causal Inference are used to construct networks of dementia from each imputed dataset. The results are then pooled into a single comprehensive network of dementia.

### 2.1 Data

The UK Biobank comprises a large cohort of over 500,000 participants aged between 40 and 69 [16]. The database contains 9,079 variables collected from 2006 to June 2023 (https://biobank.ndph.ox.ac.uk/showcase/exinfo.cgi?src=AccessingData). We utilized the ‘algorithm defined all-cause dementia’ to identify dementia cases. This definition incorporated data from baseline assessments, as well as linked data from hospital admissions and death registries [17].

To ensure a temporal order for causal inference in this prospective study design, we excluded individuals diagnosed with dementia prior to recruitment (N =177). Dementia outcomes were categorized based on the timing of diagnosis: within two years post-recruitment, more than two years post-recruitment, or none. All other variables were collected at baseline, with other disease variables only considered if they were diagnosed or reported before baseline. Our goal is to focus on modifiable risk factors for both genders. Thus, we filtered out variables related to genomics, imaging, dementia-associated health outcomes, and gender-specific health outcomes (for details, see **Supplementary Text S1**). This left 5,505 variables for further filtering (**Supplementary Table S1**).

### 2.2 Variable selection

Typically, pre-selection of dementia risk factors were either based on clinical knowledge/experience [18], or through data-driven methods like LASSO regression [19]. The first approach, while grounded in expert knowledge, can be subjective and may overlook unknown features, especially in large datasets with numerous variables, such as the UK Biobank. In contrast, the traditional data-driven methods offer more objectivity but can result in variable selection influenced by confounders or highly correlated variables, which can lead to overfitting and reduce generalizability due to dataset-specific biases [20].

Here, we novelly used pre-trained NLP models to select variable names that appeared ‘similar to’ dementia and its known risk factors [7] (Figure 1. Box2). Specifically, ‘similarity’ in this context refers to the semantic and contextual likeness that NLP models can identify between different words or phrases. Two IR models Word2Vec [14] and Doc2Vec [15] were used to understand and quantify the relationships between ‘terms’. Forty dementia-related phrases from literature [7] were used as input phrases. We used cosine similarity score to determine the similarity between the potential variable names and the target dementia-related phrases.

Four word2vec models (‘word2vec-google-news-300’, ‘glove-wiki-gigaword-300’, ‘glove-twitter-200’ and ‘fasttext-wiki-news-subwords-300’) and two doc2vec models (‘English Wikipedia DBOW’ and ‘Associated Press News DBOW’) were trained from sources such as Google News, Wikipedia and Twitter. Details of the model information can be found at https://github.com/RaRe-Technologies/gensim-data and https://github.com/jhlau/doc2vec#pre-trained-doc2vec-models. The IR models were implemented with the Genism library in Python (v 3.9). Details of the variable selection step can be found in **Supplementary Text S2**.

### 2.3 Data pre-processing and quality control

For the variables selected by IR models, we further proceeded pre-processing and multi-stage quality control (QC) (**Supplementary Text S3**). We manually reviewed these variables to assess the IR model’s performance, ensuring that any dementia risk factors not selected by IR models were added. Additionally, gender and age were included to enhance robustness. Considering chronological age is an important but non-modifiable risk factor of dementia, we also used phenotypic age as a proxy of biological age [4], [5] in parallel analysis. Unlike chronological age, phenotypic age reflects both age and disease risk [21], and importantly, is modifiable. Continuous variables were standardised by mean and standard deviation.

The sample QC filtered out individuals with mismatched gender, and related individuals, as well as the individuals with dementia diagnosis before recruitment (**Supplementary Text S4**). After these filtering steps, 406,837 participants were included in our analysis, among which 6,057 were dementia cases and 400,780 were controls (**Table 1**). Missing data in the cohort were imputed using multivariate imputation by chained equations (MICE) [22] with random forest. We set the missing rate threshold at 0.75, to ensure the retained variables exhibit a consistent pattern of missingness (**Supplementary Figure S8**). As we allowed a relatively high missing rate, the number of imputed datasets was set to 50 to well capture the original distribution of those variables with incomplete data [22]. Data imputation was performed using MICE package in R (v 4.1.0).

**Table 1.** Descriptive table of the study cohort.

| **Cohort (n = 406,781)** | **Gender Proportion (Male)** | **Chronological Age** | **Phenotypic Age** |
| --- | --- | --- | --- |
| Dementia Outcome |  | Mean (SD) | Mean (SD), NA (Missing Rate) |
| No Dementia (n = 400,730) | 54.1% | 56.4 (8.1) | 50.3 (10.3), NA = 16.7% |
| Diagnosed > 2 Years (n = 5,954) | 46.6% | 64.3 (4.7) | 60.1 (8.8), NA = 16.6% |
| Diagnosed ≤ 2 Years (n = 97) | 40.2% | 62.4 (6.9) | 62.3 (11.3), NA = 10.3% |

### 2.4 Statistical analyses

As FCI is computationally intensive [23], we firstly applied Mixed Graphical Models (MGM) [24] to rapidly infer the data structure among all selected variables. This process resulted in a skeleton structure with undirected edges between variables that represent associations without implying any directions. The skeleton structure was used as prior knowledge to inform the initial structure in the FCI algorithm [25] [25]. Detailed introduction of FCI and MGM was in **Supplementary Text S5.**

In the FCI analysis, we added an additional constraint based on the cohort’s temporal order, ensuring that dementia was not considered a cause of any other variable. Additionally, we specified that no variable could be treated as a cause of age or gender, using a ‘forbidden’ list within the FCI algorithm. False discovery rate was used to correct for multiple testing [26]. For computational efficiency, we utilized a streamlined version of FCI, known as Fast Causal Inference-Max (FCI-MAX), to deduce the final network [23]. The implementation of the analysis was performed using the ‘rCausalMGM’ package in R.

### 2.5 Results pooling from imputed datasets

In the output Partial Ancestral Graph (PAG), nodes represent variables (e.g., smoking), while edges indicate potential directional associations between them [23]. From FCI algorithm, four types of relationships between variables can be inferred from the observational data. The detailed explanation of each association/edge type inferred from the FCI algorithm can be found in **Supplementary Text S6**. To aggregate the output from each imputed dataset, we pooled results by assigning equal weights to pairwise relationships identified across datasets. Specifically, we retained associations that the FCI algorithm consistently identified, including only those present in at least 30%, 50%, or all datasets in our final model. To further refine the results, the detected structure was concentrated to highlight variables that have a direct or indirect influence on dementia onset.

## 3 Results

### 3.1 Identified causal networks of dementia and risk factors

In total, 120 variables were included in the network analysis. Our analysis identified eight variables potentially closely contributing to dementia onset (Figure 2). Among these, other mental disorders due to brain damage, dysfunctions, and to physical disease (ICD10-F06) consistently appeared to be direct contributor to dementia risks. These disorders also mediated the impact of other disorders of the brain, facial nerve disorders, and personality and behavioural disorders due to brain damage and dysfunction on dementia, with the possibility that these mediation associations might confounded by unobserved factors.

**Figure 2.**
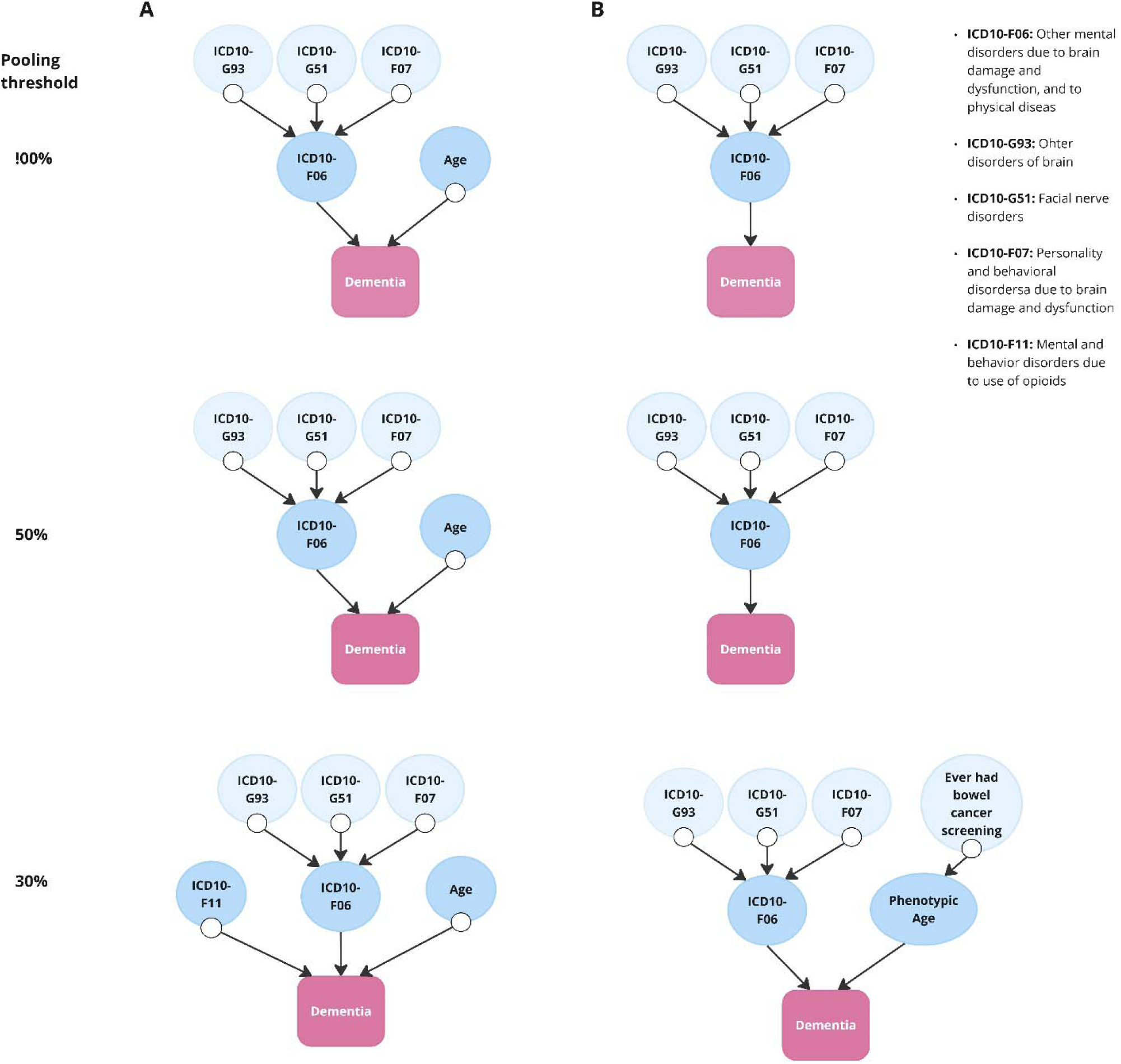
Pooled causal networks directly linked to dementia. **A.** with chronological age included **B.** with phenotypic age included instead of chronological age, using different agreement rate (pooling threshold) of 50 imputed datasets. A pooling threshold is, for each connected pair of variables, the percentage of the 50 datasets which identified the same relationships (100% -top, 50%-middle and 30%-bottom). Higher pooling threshold demonstrate more robust evidence of the associations. In each panel, the presence of an arrow “->” suggests a non-null relationship and its direction. The symbol “⍰ >” depicts three possible relationships between a pair of variables. For example, in a relationship denoted by A ⍰ > B, it is possible that 1) A directly causes B; or 2) the observed relationship between A and B is purely due to unmeasured confounding; or 3) A directly causes B, and there is unmeasured confounding between A and B.

In the network analysis including chronological age, we found strong evidence that chronological age directly affects dementia in all imputed datasets (pooling threshold = 100%). However, this relationship may be confounded by latent factors (**Figure 2A**, top panel). Additionally, there was weaker evidence (present in more than 30% but less than 50% of the imputed datasets) suggesting that mental and behaviour disorders due to opioid use may contribute to dementia risk (**Figure 2A, bottom panel**). When chronological age was replaced with phenotypic age, we found suggestive evidence that phenotypic age was directly associated with dementia, and acted as a mediator on the pathway from ever having had bowel cancer screening to dementia (**Figure 2B, bottom panel**).

Besides the pathways directly linked to dementia, consistent associations across 57 variables that might reveal interesting pathways among the diseases or risk factors related to dementia were observed (**Supplementary Table S2-S3).** A comprehensive list of the inferred pairwise relationships from each dataset is provided in **Supplementary Tables S2** and **S3**, with an explanation of variable indices in **Supplementary Table S4**.

### 3.2 Performance of variable selection using language model

The IR models selected 344 candidate variables (**Supplementary Table S5, Figure S7**), which were mapped into 24 dementia risk factor categories based on their literal meaning [7] (**Table 2**). Of the initial 40 dementia-related phrases, 10 were either not present in the UK Biobank variable list (e.g., bilingualism) or were only applicable to certain groups (e.g., hormone replacement therapy (HRT), relevant only to females) and were therefore removed from QC (**Supplementary Text S1**). Therefore, the IR model failed to identify variables related to 6 risk factors, that were “Education”, “BMI”, “Carotid atherosclerosis”, “TBI”, “hypotension” and “Anti-inflammatories”. Overall, the IR models successfully selected variables linked to 24 out of 30 manually identifiable dementia risk factors (accuracy = 0.80) from an initial pool of 5,505 variables.

**Table 2.**
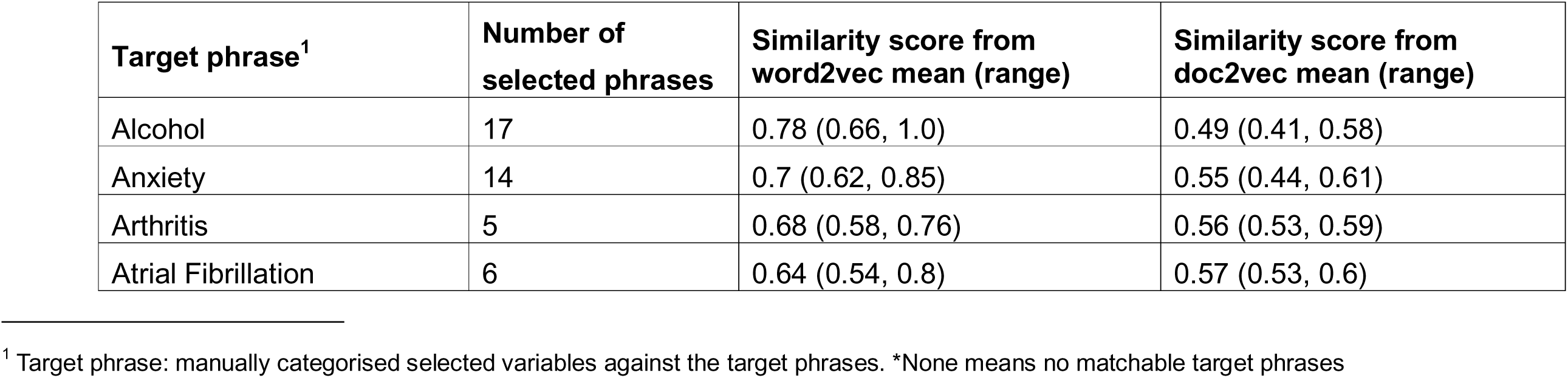
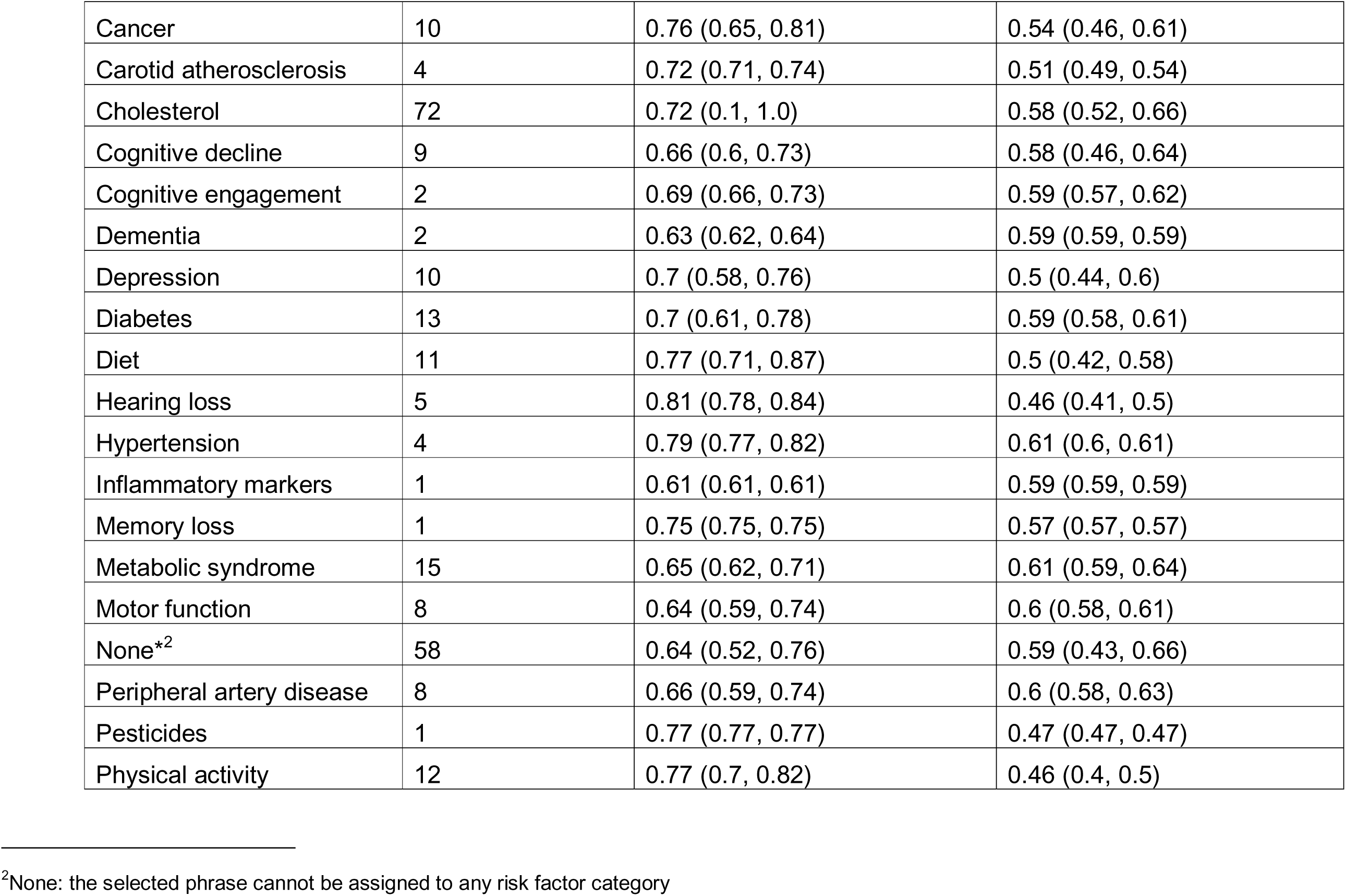
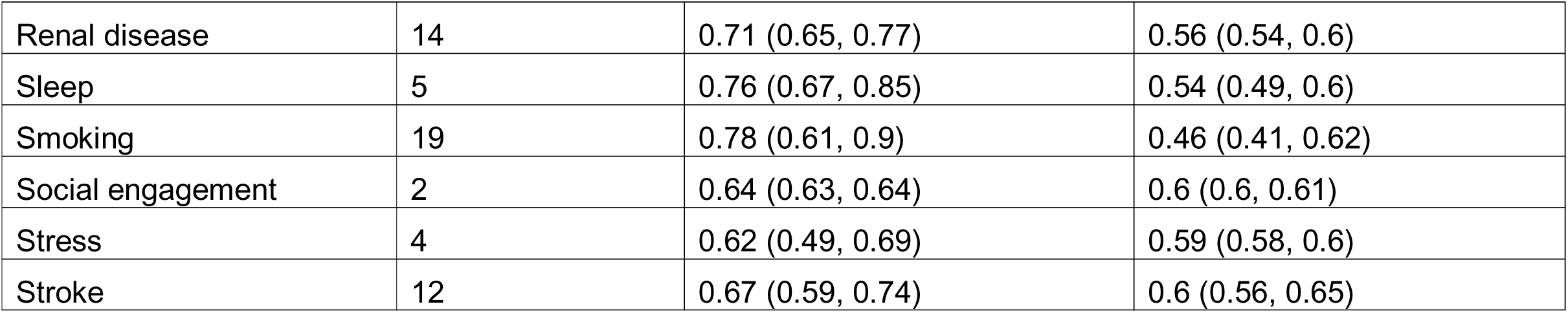
The number of variables selected by information retrieval models for each risk factor category^1^.

| **Target phrase<sup>1</sup>** | **Number of selected phrases** | **Similarity score from word2vec mean (range)** | **Similarity score from doc2vec mean (range)** |
| --- | --- | --- | --- |
| Alcohol | 17 | 0.78 (0.66, 1.0) | 0.49 (0.41, 0.58) |
| Anxiety | 14 | 0.7 (0.62, 0.85) | 0.55 (0.44, 0.61) |
| Arthritis | 5 | 0.68 (0.58, 0.76) | 0.56 (0.53, 0.59) |
| Atrial Fibrillation | 6 | 0.64 (0.54, 0.8) | 0.57 (0.53, 0.6) |

<sup>1</sup> Target phrase: manually categorised selected variables against the target phrases. \*None means no matchable target phrases

|  |  |  |  |
| --- | --- | --- | --- |
| Cancer | 10 | 0.76 (0.65, 0.81) | 0.54 (0.46, 0.61) |
| Carotid atherosclerosis | 4 | 0.72 (0.71, 0.74) | 0.51 (0.49, 0.54) |
| Cholesterol | 72 | 0.72 (0.1, 1.0) | 0.58 (0.52, 0.66) |
| Cognitive decline | 9 | 0.66 (0.6, 0.73) | 0.58 (0.46, 0.64) |
| Cognitive engagement | 2 | 0.69 (0.66, 0.73) | 0.59 (0.57, 0.62) |
| Dementia | 2 | 0.63 (0.62, 0.64) | 0.59 (0.59, 0.59) |
| Depression | 10 | 0.7 (0.58, 0.76) | 0.5 (0.44, 0.6) |
| Diabetes | 13 | 0.7 (0.61, 0.78) | 0.59 (0.58, 0.61) |
| Diet | 11 | 0.77 (0.71, 0.87) | 0.5 (0.42, 0.58) |
| Hearing loss | 5 | 0.81 (0.78, 0.84) | 0.46 (0.41, 0.5) |
| Hypertension | 4 | 0.79 (0.77, 0.82) | 0.61 (0.6, 0.61) |
| Inflammatory markers | 1 | 0.61 (0.61, 0.61) | 0.59 (0.59, 0.59) |
| Memory loss | 1 | 0.75 (0.75, 0.75) | 0.57 (0.57, 0.57) |
| Metabolic syndrome | 15 | 0.65 (0.62, 0.71) | 0.61 (0.59, 0.64) |
| Motor function | 8 | 0.64 (0.59, 0.74) | 0.6 (0.58, 0.61) |
| None*<sup>2</sup> | 58 | 0.64 (0.52, 0.76) | 0.59 (0.43, 0.66) |
| Peripheral artery disease | 8 | 0.66 (0.59, 0.74) | 0.6 (0.58, 0.63) |
| Pesticides | 1 | 0.77 (0.77, 0.77) | 0.47 (0.47, 0.47) |
| Physical activity | 12 | 0.77 (0.7, 0.82) | 0.46 (0.4, 0.5) |
| Renal disease | 14 | 0.71 (0.65, 0.77) | 0.56 (0.54, 0.6) |
| Sleep | 5 | 0.76 (0.67, 0.85) | 0.54 (0.49, 0.6) |
| Smoking | 19 | 0.78 (0.61, 0.9) | 0.46 (0.41, 0.62) |
| Social engagement | 2 | 0.64 (0.63, 0.64) | 0.6 (0.6, 0.61) |
| Stress | 4 | 0.62 (0.49, 0.69) | 0.59 (0.58, 0.6) |
| Stroke | 12 | 0.67 (0.59, 0.74) | 0.6 (0.56, 0.65) |

## 4 Discussion

### 4.1 Causal associations around nerve/brain damage disorders and dementia

We identified diseases classified under ICD10-F60, which specifically designates mental disorders resulting from brain damage and dysfunction [27], as direct contributors to dementia risk. As expected, disorders under the category may directly progress to dementia, directly increase dementia risks or share the common causal pathways with dementia. For example, mild cognitive disorder under ICD10-F60 category can progress to dementia [28]. Additionally, vascular degeneration or brain illnesses can lead to both mental disorders [29] [30] and the development of dementia [31] [32] [33], while mental disorders could also increase the risk of dementia [34].

In addition, we identified three variables --’other disorders of the brain’ (ICD10-G93), ‘facial nerve disorders’ (ICD10-G51), and ‘personality and behavioural disorders due to brain disease, damage, and dysfunctions’ (ICD10-F07), may directly affect disorders in ICD10-F60, and subsequently affect dementia, though the relationships might be confounded by unobserved factors. Each of the variable, represents a broad spectrum of disease conditions, of which many are rare. This rarity has resulted in limited literature support for some of the specific links observed in our analysis. Moreover, brain damage related to opioid use emerged as a potential, albeit weaker, contributor to dementia risk. Opioid use has been previously associated with an increased risk of dementia [35], [36], though our findings do not rule out the possibility of unobserved confounding influencing this relationship.

Nevertheless, all these variables are related to nerve or brain damage disorders, which showed the most direct connections to dementia. This suggested that neurological disorders may play a more immediate role in dementia onset compared to other risk factors and could be prioritized for targeted interventions. Furthermore, the clustering of various mental and neurological disorders indicates that these conditions may be linked to dementia through multiple pathways [37]. Additionally, interactions among these disorders may collectively contribute to the development of dementia.

### 4.2 Biological age as a potential modifiable contributor to dementia onset

Besides disorders of the nervous system, our study identified chronological age as another direct cause to dementia, with a possible presence of unobserved confounding. This is concordant with existing research findings which acknowledge that age is the known strongest risk factor for dementia [2] but not an absolute cause [38]. When age was replaced by phenotypic age —a biomarker that accounts for both chronological age and other physiological conditions, the association between dementia and phenotypic age, although less pronounced, was not confounded. Previous studies showed that clinical biomarker-based biological age was associated with the risk of dementia as well as other neurological disorders [39]. Recently developed biological age metrics derived from other omics data have also demonstrated strong performance in predicting dementia risk [40]. Our findings suggested that while chronological age itself may not have an impact on dementia, an individual’s overall health status in combination with their age may directly influence the risk of dementia. Though this marker may contain more variability and potential noise compared to absolute chronological age. More importantly, unlike chronological age, biological age is potentially modifiable through lifestyle changes, such as increased physical activity, offering valuable opportunities for dementia prevention and management. Consequently, biological ages may hold greater promise in predicting and mitigating dementia risk.

### 4.3 Other pathways around dementia risk factors

In addition, pathways identified that were not directly linked to dementia may still provide valuable biological insights (Supplementary figure S9-S10). For example, one pathway traced the progression from feelings of depression to seeking treatment and consulting a doctor (V55->V61->V20->V21). This sequence is logical, as patients often start with experiencing symptoms, followed by receiving medication and consulting healthcare providers.

While most of our findings align with existing evidence in the literature, some relationships identified in this study warrant further investigation. For example, although several studies have linked changes in total cholesterol levels to the later development of diabetes [41], [42], it remained important to validate the specific association we found between blood cholesterol levels and diabetes (V5->V40) in independent studies.

Notably, some well-established dementia risk factors [43], such as high-density lipoprotein, hearing loss, physical activity, and diabetes, appeared in the pooled causal network but were linked to dementia only indirectly, through multiple other variables such as age, metabolic rate, and sex, and potential latent factors (**Supplementary Figure 6**). This may suggest that, unlike direct contributors to dementia such as brain injury, these risk factors may influence dementia through more complex pathways, potentially making the effects of interventions on these factors less straightforward. Additionally, certain known risk factors (e.g., education [43]) did not appear in the network graph of dementia. This absence of direct paths could be due to the potential unobserved confounding and/or mediation, limited statistical power, or uncertainty introduced through data imputation.

### 4.4 Limitations in network analysis

The FCI algorithm used in our analysis is capable of identifying potential causal relationships and their directions. It uses bidirected edges to indicate potential unmeasured confounders that might affect both variables. While this approach is robust, it makes causal relationships difficult to quantify. Accurate estimation of causal effects would require further investigation with specific study designs.

By pooling results from multiple imputed datasets and setting different pooling thresholds for each pair of associations, we took into account the variation across imputed datasets and looked at the generalized overview of the networks. Here we assumed each imputed dataset contributes consistently and equally to the inference of pairwise relationships, which is difficult to test but serves as a pragmatic approach given the lack of explicit variance information or weighting criteria graphical models in the imputed datasets. Future research could benefit from enhanced pooling strategies that address both inter- and intra-group variations.

### 4.4 NLP models in variable pre-selection

The NLP models provided a novel, effective, and efficient approach for selecting candidate variables from a large pool for downstream analysis. Unlike traditional methods that rely on background knowledge [18], or standard data-driven feature selection techniques [19], we provided a new data-driven strategy that does not depend directly on the original dataset. This helps avoid common issues such as overfitting, limited power, lack of generalizability and subjective bias, allowing us to expand our exploration into previously uncharted variables and potential unknowns.

However, the application is still in its early stages, with substantial room for improvement in the future. The IR model selection accurately identified 24 out of 30 manually recognizable dementia risk factors from an initial pool of 5,505 variables. For risk factors not identified by the model, one possible reason could be variations in phrase abbreviations or formats that introduced ambiguity. During data preprocessing, terms like ‘BMI’ and ‘Anti-inflammatories’ were converted to ‘bmi’ and ‘antiinflammatories,’ leading to inconsistencies—some terms (e.g., ‘antiinflammatories’) were even challenging for human readers to interpret. Additionally, the IR model demonstrated sensitivity to formatting, with lower and upper cases represented in separate dimensions [14] [15]. These findings underscore the need to optimize preprocessing steps for improved performance.

Furthermore, the IR model occasionally introduced noise. For example, it selected phrases like ‘Carotid ultrasound authorization,’ interpreting them as similar to cardiovascular disease, reflecting limitations in contextual understanding and semantic relevance. Approximately 16.8% of the selected variables could not be classified as dementia risk factors, suggesting potential inaccuracies in the IR model. However, this discrepancy might also suggest that our understanding of dementia risk factors is incomplete; variables initially considered irrelevant could be associated with dementia through latent, unknown factors.

Finally, the accuracy of pre-trained models is significantly affected by the databases they are trained on. Some variables received very low similarity scores, likely due to their infrequent occurrence in the source databases (e.g., “avMSE”, Supplementary **Table S6 -S7**). Our models were trained on general-purpose databases like Google News and Wikipedia, which understandably differ from the specialized language used in medical research journals. Future development of models trained specifically on electronic health records [44], as well as further exploration of these approaches, could greatly enhance accuracy and relevance for medical research applications.

### 4.5 Other limitations

Due to a low prevalence of dementia in the UK Biobank, the term ‘dementia’ in our study is not subcategorised to its subtypes [45]. While subtypes like Alzheimer’s disease, vascular dementia, and Lewy body dementia share certain characteristics, they are believed to have distinct pathways [46]. Combining these conditions into the umbrella term ‘dementia’ could bring risks of overlooking the heterogeneity between subtypes, potentially leading to biases in the findings. When data collected from more dementia cases become available, it will be important to perform stratified analysis for the subtypes.

## 5. Conclusions

The identified causal network found several risk factors may closely contribute to dementia onset, offering valuable insights into the disease’s mechanisms. Beyond direct connections to brain illness, the potential direct link with biological age highlighted its possible value in dementia management. Moreover, the use of NLP models for variable selection introduced an innovative application in medical research, highlighting a promising future for advanced tools in large-scale data analyses.

## Data Availability

All data produced are available online at https://www.ukbiobank.ac.uk/enable-your-research/register

## Acknowledgements

This study used the UK Biobank application number 94719. We would like to thank the UK Biobank participants and staff for data access.

## Author contributions

XY and HG designed the study. XY performed the analyses and wrote the first draft. HG, KM and AL supervised the project. HG supervised the statistical analysis. All authors contributed to the interpretation of the results and reviewed and edited the manuscript.

## Consent statement

All human subjects provided informed consent.

## Materials and Correspondence

Correspondence to Hui Guo at or Xinzhu Yu at

## Data availability

Publicly available data from the UK Biobank study was analysed in this study. The datasets are available to researchers through an open application via https://www.ukbiobank.ac.uk/enable-your-research/register.

## Code availability

Codes in this study are available in Supplementary Text S7-S8.

